# PROTOCOL – Are clinical trial participants informed about potential benefits and harms? A comparison of informed consent materials and trial protocols

**DOI:** 10.1101/2020.05.18.20100917

**Authors:** Asger Sand Paludan-Müller, Karsten Juhl Jørgensen, Peter C. Gøtzsche

## Abstract

This is a protocol for the project entitled: “Are clinical trial participants informed about potential benefits and harms? A comparison of informed consent materials and trial protocols.”

## Background

The ‘Ethical Guidelines for Health-related Research Involving Humans’ published by the Council for International Organizations of Medical Sciences’ (CIOMS) specify that the ethical justification for such research is its scientific and social value, i.e. it must generate new knowledge that can help protect and promote health [1]. A similar sentiment is expressed in the Declaration of Helsinki, which stresses the importance of balancing risk of harms to the trial participants with the potential benefits to them and future patients [2].

One of the most important ethical requirements for clinical research involving humans is informed consent. This means, among other things, that if the participants are not adequately informed of benefits and harms, an informed decision about whether to participate in the study is not possible [1–3].

This information must be conveyed in both written and oral form. We will refer to the written material as Informed Consent Documents (ICDs).

According to the guidelines published by the Danish National Committee on Health Research Ethics, the ICD must contain information about all known or predictable harms of participating in the study and must also explicitly mention that unforeseen harms or inconveniences might occur. It is explicitly mentioned that harms must be mentioned without regard to their frequency or severity, i.e. it is not sufficient to mention only the most severe or the most common harms [4]. In the United Kingdom, the Medical Research Council has published guidance on informed consent that is less specific. It mentions that ICDs must contain “A fair and honest evaluation of the consequences of research, including possible significant benefits and harms and their relative likelihoods, must be described to potential participants.” [5]. In the United States, the Food and Drug Administration requires the ICD to contain ‘a description of any predictable risks’ as well as information on any possible discomfort and any possible benefits [6].

Thus, while exact requirements vary between countries, a description of significant known or predictable harms is universally accepted as a requirement for informed consent and the conduct of clinical research. Nonetheless, research has shown that research participants sometimes feel inadequately informed [7,8], indicating that true informed consent may sometimes not be obtained.

We will examine to what degree known benefits and harms described in unpublished clinical study protocols (CSPs) obtained from the Danish regional ethics committees match information provided to participants in ICDs. Many such trials are international multicentre trials.

## Research questions

1. Do informed consent documents provided to participants in clinical trials adequately describe known or expected benefits and harms to research participants?
2. Do informed consent documents provided to research participants in clinical trials explicitly mention that unforeseen harms might arise during the study?

## Methods

### Retrieval of trial protocols

Access to trial protocols is possible through the five regional ethics committees, which handle all applications for ethical approval of clinical trials in Denmark.

#### Inclusion criteria

- Randomised clinical trials from any clinical field

#### Exclusion criteria

- Cluster-randomised trials
- Trials with a crossover design
- Trials with only surrogate primary outcomes

We will exclude trials with only surrogate primary outcomes because determining the relevance of such outcomes to patients require detailed content area expertise from diverse clinical fields.

We will exclude trials with crossover or cluster design as these might vary systematically from parallel group trials.

#### Identification of trials

On the website of the Danish National Committee on Health Research Ethics, which functions as a common web-page for the five regional ethics committees, we will screen the titles of all research projects approved by either one of the regional ethics committees in Denmark between January 2012 and March 2013.

As the website of the Danish ethics committees only contains information on the date of approval for the trial, its title, the Danish region where the trial would take place, and the name of the prinicpal investigator, we will seek additional information about the trials through clinicaltrials.gov, the EU Clinical Trials Register, and the WHO International Clinical Trial Registry Platform [9–11]. We will use information available from the website of the ethics committee to identify the trials, e.g. the name of the intervention or a trial identifier found in the project title.

#### Preliminary assessment of eligibility

When we identify a trial in a registry, the following trial characteristics to assess eligibility will be extracted:

- Study design
- Population
- Interventions
- Inclusion and exclusion criteria
- Primary outcomes.

Eligibility will be assessed by one observer. When there is uncertainty about eligibility, a second observer will be consulted.

#### Retrieval of documents

For all trials considered potentially eligible we will send a freedom of information request to the relevant regional ethics committee. For all trials we will request copies of the following documents:

- Protocols
- Informed consent documents
- Financial agreements between study sponsors and investigators
- Publication agreements between study sponsors and investigators
- Any other relevant documents, e.g. the investigators’ brochure.

#### Final assessment of eligibility

Based on the protocols we will make a final eligibility assessment.

### Data extraction

#### Trial characteristics

One observer will extract trial characteristics from the protocols, and if necessary other documents, for all included trials. We will extract the following information:

- Title
- Medical condition studied
- Medical speciality
- Experimental intervention and comparator(s) used, including dosing schedules
- Number of arms
- Single-site or multi-centre study
- Planned sample size
- Funding source(s)
- Trial duration
- Primary outcome(s)
- Trial phase (not relevant for trials studying procedures or non-medicinal products).

The trial characteristics will be entered into an Microsoft Excel spreadsheet [12].

#### Information on benefits and harms

One observer will extract all information on benefits and harms of the active intervention from the protocols and other relevant documents (e.g. the Investigator’s Brochure if available).

From the ICD, one observer will extract the same information, i.e. all information on benefits and harms of the active intervention. We will also check whether participants are informed that unknown harms might arise.

All extracted data will be checked by a second observer.

The information on benefits and harms will be copied from the protocols (and other documents) and ICDs into a dedicated Excel spreadsheet for each trial, and all benefits and harms will be listed for both sources, to allow for comparison.

We will count the number of harms mentioned in the protocols and other documents and count how many of these that are mentioned in the ICD.

### Analysis

Using the dedicated spreadsheet, one observer will compare the information on benefits and harms provided in protocols with that in the ICDs. We will judge to what degree included studies fulfil the domains described below. All judgements will be checked by another observer. Any cases of doubt will be discussed with a senior researcher.

- Do the harms of the active intervention(s) described in the ICD match those described in the other materials available to the ethics committees?

There is some degree of subjectivity in this judgement and in cases where there is still doubt after discussion with a senior researcher, we will conservatively assume the criterion is met..

- If all harms are not described: are the harms mentioned either serious or common?

This is a subjective judgement; therefore, two researchers will independently assess whether the harms not mentioned are either serious enough or common enough that neglecting to mention them means that the information given to participants is not an honest account of the harms that can occur. In case of disagreement, a third researcher will be involved.

- Is it explicitly mentioned in the ICD that unknown harms might arise during the study?

We will only judge consider this criterion as being met if it is mentioned explicitly in the ICD. Do the benefits of the active intervention(s) described in the ICD match those described in the other materials available to the ethics committees?

As for harms, all cases of doubt will be discussed with a senior researcher, and in case of disagreement, we will conservatively assume the criterion is met.

### Statistical analyses

We will present descriptive statistics for the trial characteristics and the criteria described above.

For studies that do not mention all harms in the ICD, we will calculate the proportion of harms mentioned and will present the median and interquartile range for all studies.

We will explore potential differences between different types of funding, but as our sample size is relatively small, we will not conduct any statistical hypothesis tests.

## Strengths and limitations of the project

### Strengths of the project

To our knowledge, this will be the first comparison of descriptions of benefits and harms in materials made available for ethical approval with that in informed consent documents in a sample of recent unpublished protocols.

### Limitations of the project

This project has several limitations. Firstly, we will not compare the descriptions of harms in ICDs with harms described in the literature, only with those described in the materials made available to the research ethics committees. Additionally, even a systematic review based on journal publications might not provide exhaustive information on harms, as these are often inadequately reported [13]. Thus, It is also possible that some of the serious harms all known or expected harms are not mentioned in the clinical study protocols we will examine. CSPs and thus we might underestimate the number of ICDs not mentioning all relevant harms.

Secondly, our sample consists only of studies approved in Denmark, and results might not necessarily be applicable to other settings. However, a large proportion of studies conducted in Denmark are international multicentre trials and we are not aware of any reason that trials approved in Denmark would be systematically different from those approved in other countries.

It could be considered a limitation that our sample size will not allow us to conduct statistical hypothesis tests, e.g. to detect whether there were systematic differences between studies with different types of funding.

Finally, judgements are inherently subjective and while we will try to be conservative, it is possible that others would obtain different results based on the same materials.

## Reporting and dissemination

This protocol will be made available from the medRxiv preprint server https://www.medrxiv.org/). The results will be published in an international peer-reviewed medical journal.

## Contributions

PCG conceived the idea and wrote the study proposal. KJ and ASP contributed to the design of the study. ASP wrote the first draft of this protocol, and all authors revised the protocol and approved the final version.

## Data Availability

The data relevant to this study will be made available online on the Open Science Framework to the extent it is possible.

## References

1 Council for International Organizations of Medical Sciences (CIOMS). International Ethical Guidelines for Health-related Research Involving Humans, Fourth Edition. Geneva: 2016.

2 World Medical Association Declaration of Helsinki: Ethical Principles for Medical Research Involving Human Subjects. JAMA 2013;310:2191–4. doi:10.1001/jama.2013.281053

3 International Conference on Harmonisation. International Conference on Harmonisation of technical requirements for registration of pharmaceuticals for human use: Guideline for good clinical practice E6(R1). 1996. https://www.ich.org/fileadmin/Public_Web_Site/ICH_Products/Guidelines/Efficacy/E6/E6_R1_Guideline.pdf

4 National Videnskabsetisk Komite. At skrive en god deltagerinformation. 2011. http://www.nvk.dk/~/media/NVK/Dokumenter/At-skrive-god-deltagerinformation.pdf?la=da

5 Health Research Authority. Participant Information Sheet: What’s Involved – Consent and Participant information sheet preparation guidance. http://www.hra-decisiontools.org.uk/consent/content-sheetinvolved.html (accessed 25 Feb 2020).

6 Office of the Commissioner. Informed Consent for Clinical Trials. FDA. 2019. http://www.fda.gov/patients/clinical-trials-what-patients-need-know/informed-consent-clinicaltrials (accessed 27 Feb 2020).

7 Larson EL, Cohn EG, Meyer DD, et al. Consent administrator training to reduce disparities in research participation. J Nurs Scholarsh 2009;41:95–103. doi:10.1111/j.1547-5069.2009.01256.x

8 Koh J, Goh E, Yu K-S, et al. Discrepancy between participants’ understanding and desire to know in informed consent: are they informed about what they really want to know? J Med Ethics 2012;38:102–6. doi:10.1136/jme.2010.040972

9 US National Library of Medicine. ClinicalTrials.gov. https://clinicaltrials.gov/ (accessed 28 Feb 2020).

10 European Medicines Agency. EU Clinical Trials Register. https://www.clinicaltrialsregister.eu/ctr-search/search (accessed 28 Feb 2020).

11 World Health Organization. International Clinical Trials Registry Platform (ICTRP). http://apps.who.int/trialsearch/ (accessed 28 Feb 2020).

12 Microsoft Corporation. Microsoft Excel. 2020. https://office.microsoft.com/excel

13 Golder S, Loke YK, Wright K, et al. Reporting of Adverse Events in Published and Unpublished Studies of Health Care Interventions: A Systematic Review. PLoS Med 2016;13. doi:10.1371/journal.pmed.1002127

